# CGM glycemic persistence reflects OGTT dysglycemia

**DOI:** 10.64898/2026.04.22.26351476

**Authors:** Ren Zhang

## Abstract

**Aims:** The oral glucose tolerance test (OGTT) is effective for detecting post-load dysglycemia, but it is burdensome and therefore not routinely used. Continuous glucose monitoring (CGM) offers a convenient way to capture real-world glucose patterns, yet it remains unclear whether CGM-derived metrics reflect OGTT-defined dysglycemia. We therefore aimed to evaluate CGM-derived and clinical metrics for predicting OGTT 2-hour glucose, classifying OGTT-defined dysglycemia, and assessing day-to-day repeatability.

**Methods:** We analyzed a cohort with paired free-living CGM and OGTT. Multiple CGM-derived metrics and clinical measures were compared for prediction of OGTT 2-hour glucose, classification of OGTT-defined dysglycemia, and day-to-day stability. Predictive performance was assessed primarily by leave-one-out (LOO) R^2^, and day-to-day repeatability by intraclass correlation coefficients (ICC).

**Results:** The glycemic persistence index (GPI), a metric integrating the magnitude and duration of glycemic elevation, was the strongest single predictor of OGTT 2-hour glucose (LOO R^2^ 0.439). GPI also showed strong day-to-day repeatability (ICC 0.665) and ranked first on a combined prediction–stability score. For classification of OGTT-defined dysglycemia, HbA1c had a slightly higher AUC than GPI, but GPI plus HbA1c performed best overall, indicating complementary information.

**Conclusions:** GPI was a strong predictor of OGTT 2-hour glucose and showed a favorable balance between predictive performance and day-to-day stability, supporting its potential utility as a CGM-derived marker of dysglycemia.

## 1. Introduction

Type 2 diabetes and prediabetes are among the most common chronic metabolic conditions worldwide, and their prevalence continues to rise [1, 2]. Early identification of dysglycemia is important because glucose abnormalities often develop years before overt diabetes and are associated with increased cardiovascular and microvascular risk [3, 4]. Among reference tests, the oral glucose tolerance test (OGTT) remains a standard for characterizing post-load glucose handling, and the 2-hour glucose value in particular identifies abnormalities that fasting glucose and HbA1c do not always capture [5, 6]. In routine practice, however, the OGTT is infrequently performed because it requires fasting, a standardized glucose load, and timed venous sampling over at least two hours. As a result, a gap remains between the information that OGTT can provide and what is feasible to obtain at scale.

Continuous glucose monitoring (CGM) offers a different window into glycemia by capturing glucose patterns over multiple days under free-living conditions [7, 8]. Rather than providing a single snapshot, CGM reflects everyday glycemic exposure and fluctuation during habitual eating, activity, and sleep. As CGM use expands beyond intensive diabetes management, there is increasing interest in whether CGM-derived metrics can help characterize dysglycemia and identify individuals with abnormal glucose tolerance [9]. In cohorts with paired CGM and metabolic phenotyping, such metrics may provide a practical bridge between real-world glucose behavior and established metabolic outcomes such as OGTT 2-hour glucose, a standard diagnostic criterion for prediabetes. A key question is whether CGM-derived metrics can predict OGTT-defined dysglycemia?

A range of CGM metrics is currently available, many of which align with glycemic domains emphasized by international consensus recommendations [10, 11]. These include measures of overall glycemia, such as mean glucose and glucose management indicator (GMI); measures of glycemic variability, such as SD glucose and coefficient of variation (CV%); and range-based measures such as time in range (TIR), time above range (TAR), and time below range (TBR). Additional composite measures, including the glycemia risk index (GRI), have also been proposed. Together, these metrics provide complementary views of glycemia by emphasizing different aspects of glucose exposure and fluctuation. More recently, the glycemic persistence index (GPI) has been proposed, defined for each day as the largest integer k such that the cumulative time spent at glucose values at or above k mg/dL is at least k minutes [12]. GPI therefore integrates glycemic magnitude and duration within a single value. However, comparative evaluation of GPI alongside established CGM metrics for identifying OGTT-defined dysglycemia remains lacking.

Beyond association with a clinical outcome, another property relevant to practical use is day-to-day stability within an individual. Free-living glucose profiles vary with short-term differences in diet, meal timing, physical activity, and sleep, so repeated measurements of the same metric on different days within the same person are not expected to be identical. For a summary metric to be useful, particularly when derived from short CGM deployments, it should ideally reflect a reproducible aspect of glycemic physiology rather than be dominated by day-to-day fluctuation. Repeatability is therefore a second feature, alongside association with a clinical reference, that is informative when evaluating CGM-derived metrics.

In the present study, we evaluated various CGM-derived and clinical metrics for their ability to predict OGTT 2-hour glucose in a cohort with paired CGM and metabolic phenotyping. We assessed continuous prediction, binary classification of OGTT-defined dysglycemia, and day-to-day stability. Across the tested metrics, GPI showed the strongest overall performance and a favorable balance between prediction and repeatability, supporting its potential utility as a CGM-derived marker of dysglycemia.

## 2. Materials and Methods

### 2.1. Study cohort and data source

We analyzed publicly available data the study of free-living CGM and metabolic phenotyping in adults[13]. The dataset included Dexcom G4 CGM recordings together with standardized 75-g oral glucose tolerance test (OGTT) measurements, fasting glucose, HbA1c, fasting insulin, body mass index (BMI), and steady-state plasma glucose (SSPG). The original study enrolled 57 participants and performed CGM during usual daily life in parallel with metabolic testing.

### 2.2. CGM preprocessing

Glucose values were grouped by participant and calendar day. Non-numeric low-sensor flags were mapped to 40 mg/dL. Days with fewer than 200 valid readings were excluded. Daily CGM metrics were first calculated within each qualifying day and then averaged across days within each participant to generate subject-level values for prediction analyses. For repeatability analyses, daily metric values were retained to quantify within-subject day-to-day stability.

### 2.3. CGM and clinical metrics

We evaluated 17 candidate metrics for prediction of OGTT 2-hour glucose, including CGM-derived metrics and clinical laboratory comparators. The CGM panel, according to international consensus recommendations for CGM reporting [11], included mean glucose, SD glucose, coefficient of variation (CV%), glucose management indicator (GMI), glycemia risk index (GRI), time in range (TIR 70–180%), tight range (70–140%), time below range (TBR <70% and <54%), time above range (TAR >180%, 181–250%, and >250%), and the glycemic persistence index (GPI), a recently proposed metric intended to summarize the magnitude and persistence of glycemic elevation in a single value. Mean glucose and SD glucose were calculated directly from daily CGM values. CV% was calculated as 100 × SD / mean glucose. GMI was calculated from mean glucose using the published linear conversion. GRI was calculated from standard low- and high-glucose exposure bins. TIR metrics were calculated as the percentage of CGM readings within the specified glucose intervals, and TAR/TBR metrics were calculated as the percentage of readings above or below the stated thresholds. The non-CGM comparator panel included HbA1c, fasting glucose, fasting insulin, BMI, and SSPG. The CGM metrics were selected to include widely used and consensus-recommended summary measures together with GPI as the persistence metric of interest.

### 2.4. Definition of GPI

GPI was calculated from each day of CGM data as the largest integer k such that the cumulative duration spent at glucose values greater than or equal to k mg/dL was at least k minutes during that day [12]. Daily GPI values were averaged across qualifying days within each participant for subject-level prediction analyses.

### 2.5. Continuous prediction analyses

The primary continuous outcome was OGTT 2-hour glucose. For each metric, predictive performance was summarized using Pearson correlation, ordinary least-squares (OLS) R^2^, and leave-one-out (LOO) R^2^. Pearson correlation and OLS *R*^2^ describe in-sample association. LOO R^2^ was used as the primary ranking metric because it evaluates out-of-sample prediction. For each participant, a univariate linear model was fit in all remaining participants and then used to predict the held-out participant’s OGTT 2-hour glucose. This process was repeated for every participant, and LOO R^2^ was calculated from the full set of held-out predictions.

### 2.6. Day-to-day repeatability analyses

Repeatability analyses were restricted to participants with at least 6 qualifying CGM days. Day-to-day repeatability was assessed using the intraclass correlation coefficient (ICC) and the median within-subject coefficient of variation (CV). ICC was used as the primary repeatability measure because it directly assesses whether a metric is dominated by stable between-person differences rather than within-person day-to-day fluctuation. In contrast, median within-subject CV is scale-dependent and therefore not ideal for comparing repeatability across metrics with different units, ranges, or transformations. CV was therefore treated as a complementary descriptive measure rather than the main basis for cross-metric repeatability comparisons. Ninety-five percent confidence intervals for ICC were estimated by subject-level bootstrap resampling.

### 2.7. Prediction-stability score

To summarize the combined utility of each metric, we defined a prediction-stability score (PSS) as:

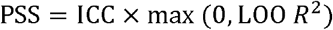

Higher PSS values indicate a more favorable balance between predictive performance and day-to-day repeatability.

### 2.8. Binary classification analyses

For binary analyses, OGTT-defined dysglycemia was defined as OGTT 2-hour glucose ≥140 mg/dL. Binary discrimination was evaluated using receiver operating characteristic (ROC) analysis and leave-one-out area under the curve (LOO AUC). For each model, predicted probabilities were generated in a leave-one-out framework using logistic regression, and the resulting held-out predictions were used to construct ROC curves and calculate AUC.

### 2.9. Decision-rule analyses

To translate continuous GPI values into a candidate decision rule, we evaluated sensitivity, specificity, and the Youden index across candidate GPI thresholds for identifying OGTT-defined dysglycemia. The Youden index was defined as sensitivity + specificity − 1. Based on this analysis, a rounded GPI threshold of 135 was selected for rule-based comparisons. We then evaluated GPI ≥135, HbA1c ≥5.7%, GPI ≥135 OR HbA1c ≥5.7%, and GPI ≥135 AND HbA1c ≥5.7%. For each rule, we calculated sensitivity, specificity, positive predictive value (PPV), negative predictive value (NPV), and the Youden index.

### 2.10. Statistical analysis

All analyses were performed at the participant level unless otherwise stated. Continuous prediction analyses used subject-level averages of daily CGM metrics. Repeatability analyses used day-level values. *p* values were from two-sided Pearson correlation tests with OGTT 2-hour glucose. Two-sided *p* values <0.05 were considered statistically significant. All primary data processing and statistical analyses were performed in Python using custom scripts together with pandas, NumPy, SciPy, scikit-learn, and matplotlib; figures were generated from these analysis outputs with final formatting as needed in Origin (OriginLab, Northampton, MA, USA).

## 3. Results

### 3.1. GPI was the strongest single CGM metric for continuous prediction of OGTT 2-hour glucose

Analyses were performed in the study cohort with available CGM and standardized 75-g oral glucose tolerance test (OGTT) data. Continuous prediction analyses included 54 participants with both CGM and OGTT measurements; HbA1c was available in 53. We compared GPI with 17 candidate metrics, including consensus-recommended CGM summaries and routine clinical laboratory measures, for prediction of OGTT 2-hour glucose (Table 1). Performance was summarized by Pearson correlation, ordinary least-squares (OLS) R^2^, and leave-one-out (LOO) R^2^. Pearson correlation and OLS R^2^ describe in-sample association. By contrast, LOO R^2^ was used as the primary measure of predictive performance because it evaluates out-of-sample prediction: for each participant, the model was fit in all remaining participants and then used to predict the held-out participant’s OGTT 2-hour glucose. This process was repeated for every participant, and the resulting LOO R^2^ therefore reflects how well each metric generalized beyond the data used to fit the model. We report LOO R^2^ as the primary measure of predictive accuracy, with OLS R^2^ and the Pearson correlation coefficient (*r*) provided alongside to describe the in-sample association and its strength, respectively.

**Table 1.**
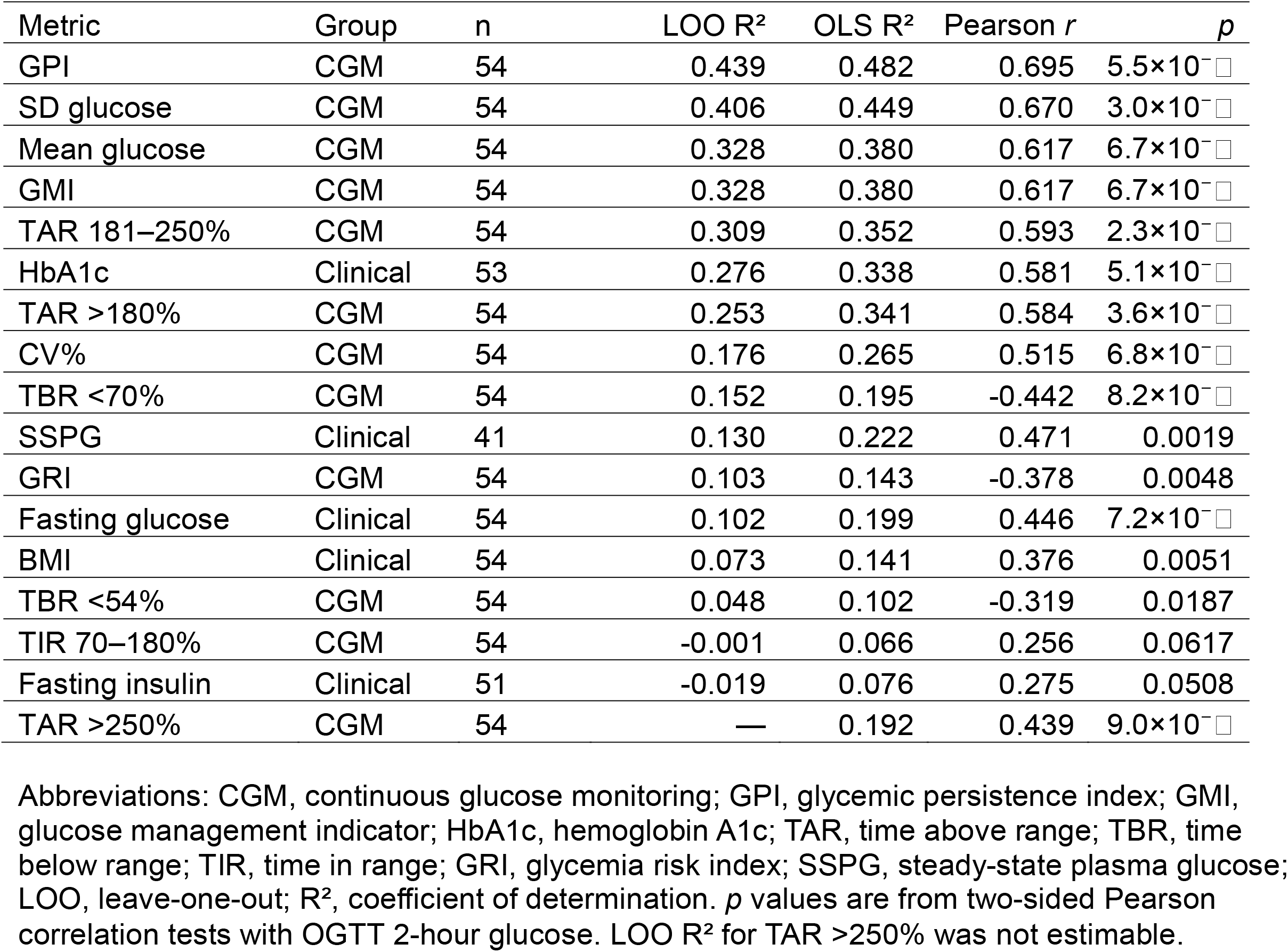
Continuous prediction of OGTT 2-hour glucose.

Among all tested metrics, GPI showed the strongest overall performance by the primary ranking metric, LOO R^2^, with a value of 0.439 (Pearson r 0.695, OLS R^2^ 0.482; Table 1). The closest competing CGM metric was SD glucose (LOO R^2^ 0.406, Pearson r 0.670, OLS R^2^ 0.449), whereas mean glucose and GMI were weaker but still informative, each with LOO R^2^ 0.328 (Pearson r 0.617, OLS R^2^ 0.380). Threshold-based hyperglycemia metrics showed intermediate performance, including TAR 181–250% (LOO R^2^ 0.309) and TAR >180% (LOO R^2^ 0.253).

Among non-CGM clinical markers, HbA1c was the strongest comparator (Pearson r 0.581, OLS R^2^ 0.338, LOO R^2^ 0.276), but it remained inferior to GPI. Fasting glucose performed more weakly (Pearson r 0.446, OLS R^2^ 0.199, LOO R^2^ 0.102), and fasting insulin performed poorly (Pearson r 0.275, OLS R^2^ 0.076, LOO R^2^ −0.019). Together, these results identify GPI as the leading single-metric predictor of OGTT 2-hour glucose in this cohort.

### 3.2. GPI provided a favorable overall balance between prediction and day-to-day repeatability

For CGM-derived summary metrics, day-to-day stability is especially important because free-living glucose profiles vary with short-term differences in diet, meal timing, physical activity, sleep, and other daily behaviors. A useful metric should therefore reflect a reproducible aspect of glycemic physiology rather than short-term noise. This is particularly relevant for short-duration CGM deployments, in which unstable metrics may show apparent associations yet perform inconsistently across days. We therefore evaluated day-to-day repeatability, in addition to continuous prediction of OGTT 2-hour glucose, to identify metrics that combined predictive value with practical stability.

Day-to-day repeatability was assessed using the intraclass correlation coefficient (ICC) and median within-subject coefficient of variation (CV). ICC was used as the primary repeatability measure because it directly assesses whether a metric is dominated by stable between-person differences rather than within-person day-to-day fluctuation. In contrast, median within-subject CV is scale-dependent and therefore not ideal for comparing repeatability across metrics with different units, ranges, or transformations. CV was therefore treated as a complementary descriptive measure, whereas ICC was used as the main basis for cross-metric repeatability comparisons. To integrate repeatability with predictive performance, we defined a prediction– stability score (PSS) as ICC × max(0, leave-one-out R^2^), with higher values indicating a better combined balance of stability and prediction.

Among the tested CGM metrics, mean glucose and GMI showed the highest repeatability, each with ICC 0.711 (95% CI 0.576–0.816; Table 2). GPI also showed strong day-to-day repeatability, with ICC 0.665 (95% CI 0.522–0.786) and median within-subject CV 7.897%. SD glucose was less repeatable (ICC 0.507, 95% CI 0.316–0.665; median within-subject CV 23.559%), and most threshold-based metrics showed lower repeatability and wider uncertainty (Figure 1A). When repeatability and predictive performance were combined using the prediction–stability score (PSS), GPI ranked first (0.292), followed by mean glucose (0.233), GMI (0.233), and SD glucose (0.206) (Figure 1B; Table 2). Thus, although mean glucose and GMI were slightly more repeatable, GPI provided the best overall balance between OGTT prediction and day-to-day stability in this cohort.

**Table 2.** Stability and combined utility.

| Metric | PSS | ICC | 95% CI for ICC | Median within-subject CV (%) |
| --- | --- | --- | --- | --- |
| GPI | 0.292 | 0.665 | 0.522–0.786 | 7.897 |
| Mean glucose | 0.233 | 0.711 | 0.576–0.816 | 6.324 |
| GMI | 0.233 | 0.711 | 0.576–0.816 | 2.727 |
| SD glucose | 0.206 | 0.507 | 0.316–0.665 | 23.559 |
| TAR 181–250% | 0.108 | 0.351 | 0.108–0.559 | 0.000 |
| TAR >180% | 0.106 | 0.418 | 0.198–0.604 | 0.000 |
| CV% | 0.074 | 0.418 | 0.233–0.582 | 22.330 |
| TBR <70% | 0.061 | 0.399 | 0.199–0.572 | 115.608 |
| GRI | 0.039 | 0.383 | 0.197–0.548 | 108.030 |
| TBR <54% | 0.005 | 0.097 | -0.059–0.278 | 0.000 |
| TIR 70–180% | 0.000 | 0.353 | 0.162–0.539 | 3.356 |
| Tight range 70–140% | 0.000 | 0.291 | 0.064–0.478 | 7.500 |
| TAR >250% | 0.000 | 0.147 | 0.000–0.332 | 0.000 |

Abbreviations: PSS, prediction–stability score; ICC, intraclass correlation coefficient; CV, coefficient of variation; GPI, glycemic persistence index; GMI, glucose management indicator; TAR, time above range; TBR, time below range; TIR, time in range; GRI, glycemia risk index. Stability analyses were restricted to 50 subjects with at least 6 qualifying CGM days ( $\geq 200$ readings/day). Ninety-five percent confidence intervals for ICC were estimated by subject-level bootstrap. PSS was defined as $ICC \times \max(0, LOO R^2)$ .

**Figure 1.**
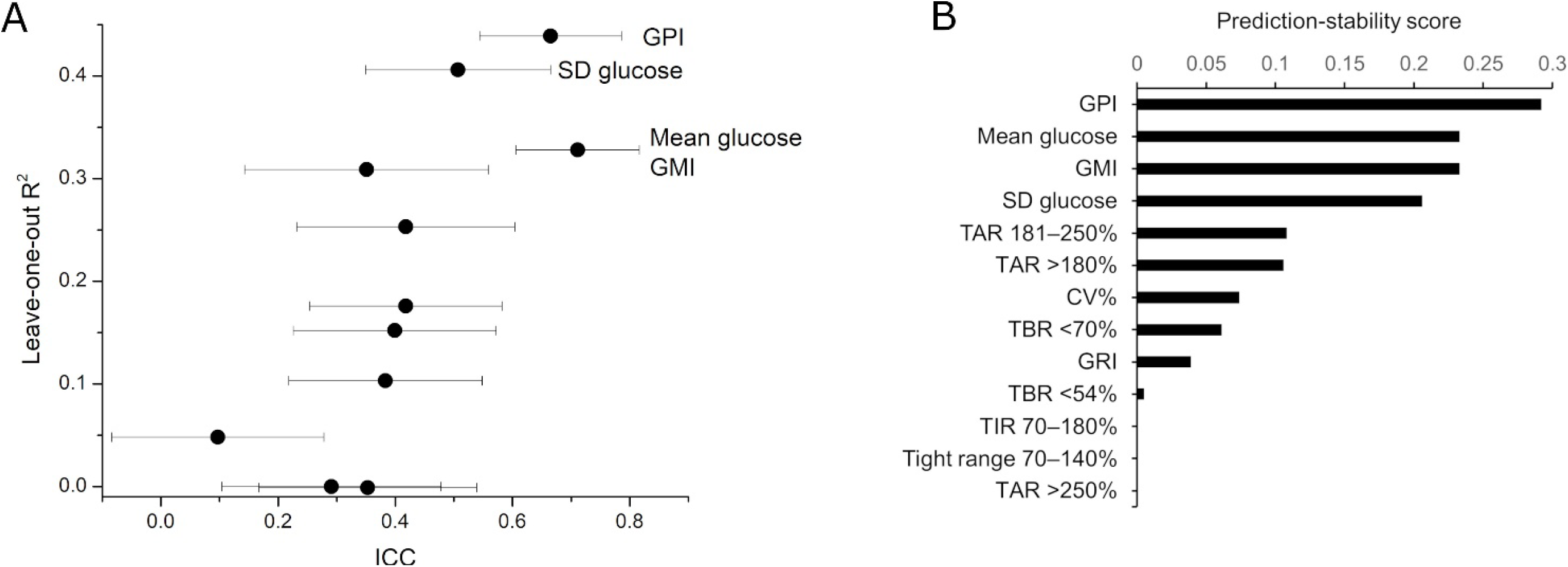
Performance of CGM metrics in predicting OGTT dysglycemia. A) Leave-one-out R^2^ and ICC across CGM metrics. Leave-one-out (LOO) R^2^ and intraclass correlation coefficient (ICC) across CGM metrics. The x-axis shows ICC, with horizontal error bars indicating 95% confidence intervals, and the y-axis shows leave-one-out R^2^ for prediction of OGTT 2-hour glucose. Metrics located toward the upper-right combine stronger predictive performance with greater day-to-day repeatability. Top-performing metrics are labeled for clarity, whereas the remaining tested CGM metrics are included as unlabeled points to reduce. Unlabeled metrics include TBR <70%, TBR <54%, TIR 70–180%, and tight range 70–140%. B) Prediction–stability score (PSS) ranking of CGM metrics. PSS was defined as ICC × max(0, leave-one-out R^2^), with higher values indicating a better overall balance between repeatability and predictive performance. GPI ranked first, followed by mean glucose, GMI, and SD glucose.

### 3.3. GPI and HbA1c provided complementary information for classifying OGTT-defined dysglycemia

We next evaluated binary classification of OGTT-defined dysglycemia, defined as OGTT 2-hour glucose ≥140 mg/dL, using receiver operating characteristic (ROC) analysis (Figure 2A; Table 3). In this setting, HbA1c slightly outperformed GPI alone, with a leave-one-out area under the curve (AUC) of 0.839 versus 0.825 for GPI. However, GPI remained superior to other single CGM metrics, including mean glucose (AUC 0.808), GMI (AUC 0.783), and SD glucose (AUC 0.724). Thus, although HbA1c was the strongest single marker for binary classification, GPI remained the best-performing single CGM-derived metric in this analysis.

**Table 3.** ROC analysis for OGTT 2-hour glucose ≥140 mg/dL.

| Model | Group | n | LOO AUC |
| --- | --- | --- | --- |
| GPI + HbA1c | Combined | 55 | 0.861 |
| HbA1c | Clinical | 55 | 0.839 |
| GPI | CGM | 57 | 0.825 |
| Fasting glucose | Clinical | 56 | 0.816 |
| Mean glucose | CGM | 57 | 0.808 |
| GMI | CGM | 57 | 0.783 |
| SD glucose | CGM | 57 | 0.724 |

Abbreviations: GPI, glycemic persistence index; GMI, glucose management indicator; HbA1c, hemoglobin A1c; LOO, leave-one-out; AUC, area under the receiver operating characteristic curve.

**Figure 2.**
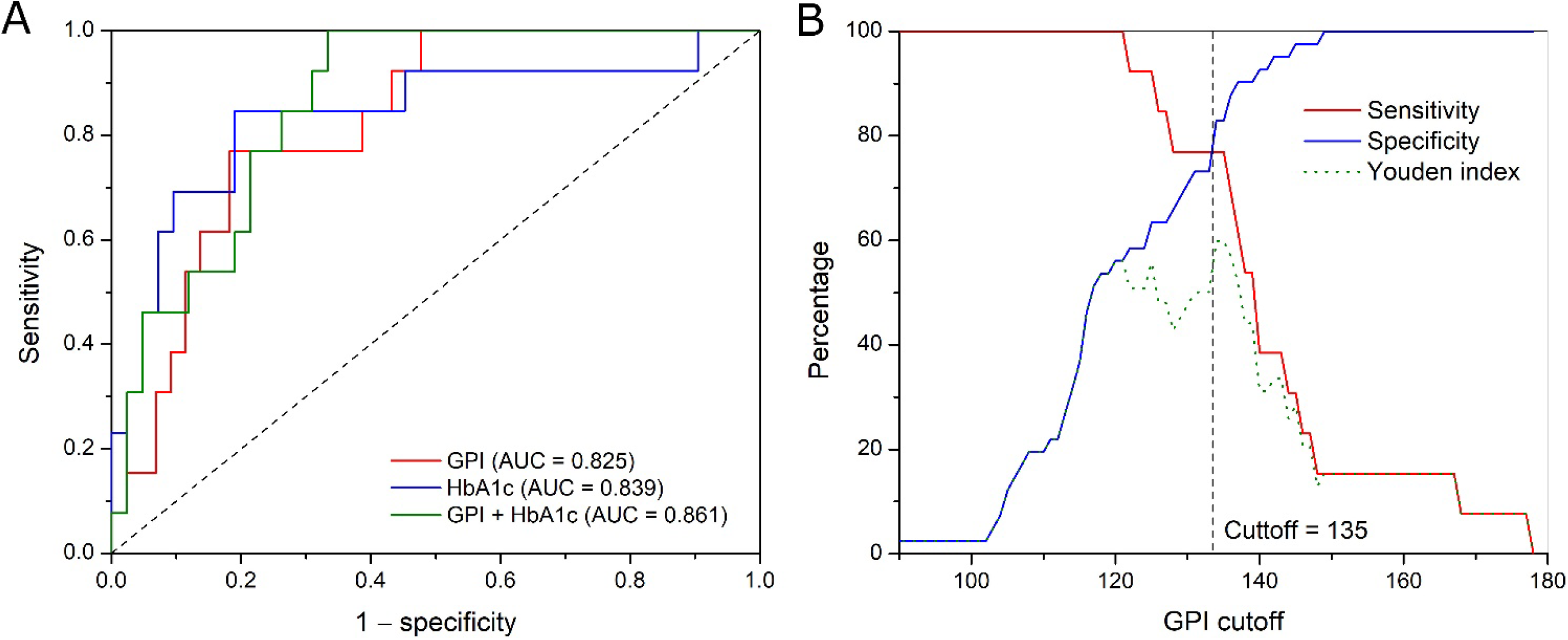
Binary classification of OGTT-defined dysglycemia and exploratory GPI cutoff analysis. A) Receiver operating characteristic curves for classification of OGTT 2-hour glucose ≥140 mg/dL using GPI, HbA1c, and the combination of GPI plus HbA1c. The combined model showed the highest area under the curve, indicating complementary information from CGM-derived glycemic persistence and laboratory glycemia. B) Sensitivity, specificity, and Youden index across candidate GPI cutoffs for identifying OGTT 2-hour glucose ≥140 mg/dL. The vertical dashed line indicates the rounded cutoff of GPI = 135 used for decision-rule analyses.

Combining GPI and HbA1c produced the highest overall discriminative performance, with a leave-one-out AUC of 0.861. This result indicates that CGM-derived glycemic persistence and laboratory glycemia capture overlapping but non-identical aspects of dysglycemia, and that their joint use improves classification of OGTT-defined abnormality. Together, these results support a complementary rather than substitutive role for GPI alongside HbA1c in identifying OGTT-defined dysglycemia.

### 3.4. Decision-rule performance of a GPI cutoff of 135, alone and combined with HbA1c

To evaluate rule-based classification of OGTT-defined dysglycemia (2-hour glucose ≥140 mg/dL), we examined sensitivity, specificity, and the Youden index across candidate GPI thresholds (Figure 2B). Using the Youden index, the cutoff was determined to be GPI = 135. This threshold should be regarded as exploratory and cohort-specific. Applied alone, GPI ≥135 yielded sensitivity 0.77, specificity 0.82, PPV 0.56, NPV 0.92, and a Youden index of 0.59 (n = 57; Table 4). For comparison, HbA1c ≥5.7% yielded sensitivity 0.62, specificity 0.93, PPV 0.73, NPV 0.89, and a Youden index of 0.54 (n = 55). Thus, GPI ≥135 was more sensitive, whereas HbA1c ≥5.7% was more specific.

**Table 4.** Decision-rule performance for OGTT 2-hour glucose ≥140 mg/dL.

| Rule | n | Sensitivity | Specificity | PPV | NPV | Youden index |
| --- | --- | --- | --- | --- | --- | --- |
| GPI $\geq 135$ | 57 | 0.77 | 0.82 | 0.56 | 0.92 | 0.59 |
| HbA1c $\geq 5.7\%$ | 55 | 0.62 | 0.93 | 0.73 | 0.89 | 0.54 |
| GPI $\geq 135$ OR HbA1c $\geq 5.7\%$ | 55 | 1.00 | 0.79 | 0.59 | 1.00 | 0.79 |
| GPI $\geq 135$ AND HbA1c $\geq 5.7\%$ | 55 | 0.38 | 0.95 | 0.71 | 0.83 | 0.34 |

Abbreviations: GPI, glycemic persistence index; HbA1c, hemoglobin A1c; PPV, positive predictive value; NPV, negative predictive value. Binary outcome was OGTT 2-hour glucose $\geq 140$ mg/dL. Sample size differed across rules because HbA1c was unavailable for two participants. Youden index was defined as sensitivity + specificity – 1.

Because GPI and HbA1c provided complementary information, we next evaluated combined rules. The OR rule (GPI ≥135 or HbA1c ≥5.7%) achieved sensitivity 1.00, specificity 0.79, PPV 0.59, NPV 1.00, and the highest Youden index (0.79; n = 55). By contrast, the AND rule (GPI ≥135 and HbA1c ≥5.7%) achieved sensitivity 0.38, specificity 0.95, PPV 0.71, NPV 0.83, and a Youden index of 0.34. Together, these results support an exploratory role for GPI ≥135, alone or combined with HbA1c, in rule-based identification of OGTT-defined dysglycemia.

## 4. Discussion

As continuous glucose monitoring is increasingly used to characterize glycemia beyond traditional diabetes management, there is growing interest in which CGM-derived metrics most meaningfully reflect underlying dysglycemia [8, 14]. In this study, we compared multiple CGM-derived and clinical metrics against OGTT-based measures and found that GPI showed the strongest overall performance, particularly for continuous prediction of OGTT 2-hour glucose and for balancing predictive value with day-to-day repeatability. For binary classification of OGTT-defined dysglycemia, HbA1c slightly outperformed GPI alone, but GPI remained the best-performing single CGM-derived metric, and the combination of GPI and HbA1c achieved the highest overall discrimination.

GPI may stand out among CGM metrics in predicting OGTT glucose because it captures a feature of glycemia that is relevant to the physiological abnormality reflected by OGTT 2-hour glucose. By integrating both the magnitude and duration of glycemic elevation within a single value, GPI may better reflect sustained glycemic exposure during daily life than metrics that emphasize average glucose, glycemic variability, or time spent above a fixed threshold alone[12]. This interpretation is consistent with the present findings, although the physiological basis for GPI’s performance will require further validation in independent cohorts and mechanistic studies.

GPI and HbA1c summarize glycemia on different timescales and through different biological processes. HbA1c reflects average glucose exposure over the preceding two to three months through nonenzymatic hemoglobin glycation, whereas GPI summarizes the magnitude and persistence of glucose elevations observed directly during short periods of free-living monitoring. Consistent with this distinction, the combination of GPI and HbA1c outperformed either marker alone for classification of OGTT-defined dysglycemia, indicating that the two provide partially non-overlapping information. These findings support a complementary rather than substitutive role for GPI alongside HbA1c and suggest that CGM-derived persistence metrics may add useful information beyond established laboratory measures.

From a translational perspective, the decision-rule analysis provides an initial illustration of how GPI might be used in practice. The exploratory cutoff selected by the Youden index was GPI = 135, corresponding to at least 135 minutes per day with glucose ≥135 mg/dL. As a stand-alone rule, this threshold showed a reasonable balance of sensitivity and specificity for identifying OGTT-defined dysglycemia. When combined with HbA1c using an OR rule, performance improved further, with sensitivity reaching 100% in this cohort at the cost of some reduction in specificity. Although these findings are preliminary and cohort-specific, they suggest that GPI may be useful not only as a continuous metric but also as the basis for clinically interpretable screening-style rules.

Day-to-day stability is an important property of CGM-derived metrics, particularly when they are derived from short free-living recordings in which day-level variation in diet, physical activity, sleep, and related behaviors can introduce substantial fluctuation. In this cohort, GPI showed day-to-day repeatability comparable to mean glucose and GMI and higher than most threshold-based metrics, indicating that a meaningful proportion of its variation reflects stable between-person differences rather than within-person day-to-day noise. This stability supports the feasibility of summarizing GPI from relatively short CGM deployments. More broadly, the prediction–stability score framework illustrates that CGM metrics are most informatively evaluated not only by their association with a reference outcome, but also by their reproducibility across days.

This study has both strengths and limitations. A strength of this study is the head-to-head evaluation of multiple CGM-derived and clinical metrics in a cohort with paired free-living CGM and standardized metabolic phenotyping, assessing both predictive performance and day-to-day repeatability rather than prediction alone. Several limitations should be considered. First, the sample size was modest, which may limit precision and affect the stability of metric ranking. Second, this was a single-cohort analysis, and external validation in independent populations will be necessary to determine generalizability. Third, although OGTT 2-hour glucose is a clinically relevant reference measure, free-living CGM and OGTT reflect related but not identical aspects of glucose physiology. Finally, the GPI cutoff of 135 should be regarded as exploratory pending external validation.

These limitations also point to several directions for future work. Validation in larger and independent cohorts will be important to determine the generalizability of the present findings and the robustness of metric ranking across populations. It will also be useful to evaluate GPI across broader demographic and metabolic groups, different CGM devices, and varying monitoring durations and preprocessing choices. In addition, future studies should test whether GPI adds value beyond established laboratory and CGM measures in multivariable models and whether it predicts prospective outcomes, including progression to diabetes or other clinically relevant metabolic endpoints.

## 5. Conclusion

In summary, among the CGM-derived metrics evaluated, GPI showed the strongest performance for continuous prediction of OGTT 2-hour glucose and a favorable balance between predictive value and day-to-day stability. GPI also complemented HbA1c for classification of OGTT-defined dysglycemia, suggesting that these measures capture partially distinct aspects of glycemia. These findings support GPI as a promising CGM-derived marker of dysglycemia. Further validation in larger and independent cohorts is warranted.

## Author Contributions

R.Z. conceptualized, designed the study, analyzed the data, wrote the paper, and obtained the funding.

## Data Availability Statement

The CGM dataset analyzed in this study is publicly available. The Python scripts used for analysis are available from the corresponding author upon reasonable request.

## References

[1] Abdul Basith Khan M, Hashim MJ, King JK, Govender RD, Mustafa H, Al Kaabi J. Epidemiology of type 2 diabetes—global burden of disease and forecasted trends. Journal of epidemiology and global health. 2020;10:107–11.

[2] Rooney MR, Fang M, Ogurtsova K, Ozkan B, Echouffo-Tcheugui JB, Boyko EJ, et al. Global prevalence of prediabetes. Diabetes Care. 2023;46:1388–94.

[3] Tabák AG, Herder C, Rathmann W, Brunner EJ, Kivimäki M. Prediabetes: a high-risk state for diabetes development. The Lancet. 2012;379:2279–90.

[4] Echouffo-Tcheugui JB, Selvin E. Prediabetes and what it means: the epidemiological evidence. Annu Rev Public Health. 2021;42:59–77.

[5] Jagannathan R, Neves JS, Dorcely B, Chung ST, Tamura K, Rhee M, et al. The oral glucose tolerance test: 100 years later. Diabetes Metab Syndr Obes. 2020:3787–805.

[6] Bartoli E, Fra G, Schianca GC. The oral glucose tolerance test (OGTT) revisited. Eur J Intern Med. 2011;22:8–12.

[7] Rodbard D. Continuous glucose monitoring: a review of successes, challenges, and opportunities. Diabetes Technol Ther. 2016;18:S2-3-S2-13.

[8] Klonoff DC, Ahn D, Drincic A. Continuous glucose monitoring: a review of the technology and clinical use. Diabetes Res Clin Pract. 2017;133:178–92.

[9] Keshet A, Shilo S, Godneva A, Talmor-Barkan Y, Aviv Y, Segal E, et al. CGMap: characterizing continuous glucose monitor data in thousands of non-diabetic individuals. Cell metabolism. 2023;35:758–69. e3.

[10] Kovatchev BP. Metrics for glycaemic control—from HbA1c to continuous glucose monitoring. Nature Reviews Endocrinology. 2017;13:425–36.

[11] Battelino T, Alexander CM, Amiel SA, Arreaza-Rubin G, Beck RW, Bergenstal RM, et al. Continuous glucose monitoring and metrics for clinical trials: an international consensus statement. The lancet Diabetes & endocrinology. 2023;11:42–57.

[12] Zhang R. Defining a glycemic persistence index (GPI) for continuous glucose monitoring. [Preprint] Research Square. 2026: 10.21203/rs.3.rs-8841862/v2.

[13] Hall H, Perelman D, Breschi A, Limcaoco P, Kellogg R, McLaughlin T, et al. Glucotypes reveal new patterns of glucose dysregulation. PLoS Biol. 2018;16:e2005143.

[14] Bender C, Vestergaard P, Cichosz SL. The history, evolution and future of Continuous Glucose Monitoring (CGM). Diabetology. 2025;6:17.

